# Effect of Montelukast vs Placebo on Time to Sustained Recovery in Outpatients with COVID-19: The ACTIV-6 Randomized Clinical Trial

**DOI:** 10.1101/2024.05.16.24307115

**Authors:** Russell L. Rothman, Thomas G. Stewart, Ahmad Mourad, David R. Boulware, Matthew W. McCarthy, Florence Thicklin, Idania T. Garcia del Sol, Jose Luis Garcia, Carolyn T. Bramante, Nirav S. Shah, Upinder Singh, John C. Williamson, Paulina A. Rebolledo, Prasanna Jagannathan, Tiffany Schwasinger-Schmidt, Adit A. Ginde, Mario Castro, Dushyantha Jayaweera, Mark Sulkowski, Nina Gentile, Kathleen McTigue, G. Michael Felker, Allison DeLong, Rhonda Wilder, Sean Collins, Sarah E. Dunsmore, Stacey J. Adam, George J. Hanna, Elizabeth Shenkman, Adrian F. Hernandez, Susanna Naggie, Christopher J. Lindsell, Accelerating COVID-19 Therapeutic Interventions and Vaccines (ACTIV)-6 Study Group and Investigators

## Abstract

**Importance:** The effect of montelukast in reducing symptom duration among outpatients with mild to moderate coronavirus disease 2019 (COVID-19) is uncertain.

**Objective:** To assess the effectiveness of montelukast compared with placebo in treating outpatients with mild to moderate COVID-19.

**Design, Setting, and Participants:** The ACTIV-6 platform randomized clinical trial aims to evaluate the effectiveness of repurposed medications in treating mild to moderate COVID-19. Between January 27, 2023, and June 23, 2023, 1250 participants ≥30 years of age with confirmed SARS-CoV-2 infection and ≥2 acute COVID-19 symptoms for ≤7 days, were included across 104 US sites to evaluate the use of montelukast.

**Interventions:** Participants were randomized to receive montelukast 10 mg once daily or matched placebo for 14 days.

**Main Outcomes and Measures:** The primary outcome was time to sustained recovery (defined as at least 3 consecutive days without symptoms). Secondary outcomes included time to death; time to hospitalization or death; a composite of hospitalization, urgent care visit, emergency department visit, or death; COVID clinical progression scale; and difference in mean time unwell.

**Results:** Among participants who were randomized and received study drug, the median age was 53 years (IQR 42–62), 60.2% were female, 64.6% identified as Hispanic/Latino, and 56.3% reported ≥2 doses of a SARS-CoV-2 vaccine. Among 628 participants who received montelukast and 622 who received placebo, differences in time to sustained recovery were not observed (adjusted hazard ratio [HR] 1.02; 95% credible interval [CrI] 0.92–1.12; P(efficacy) = 0.63]). Unadjusted median time to sustained recovery was 10 days (95% confidence interval 10–11) in both groups. No deaths were reported and 2 hospitalizations were reported in each group; 36 participants reported healthcare utilization events (a priori defined as death, hospitalization, emergency department/urgent care visit); 18 in the montelukast group compared with 18 in the placebo group (HR 1.01; 95% CrI 0.45–1.84; P(efficacy)=0.48). Five participants experienced serious adverse events (3 with montelukast and 2 with placebo).

**Conclusions and Relevance:** Among outpatients with mild to moderate COVID-19, treatment with montelukast does not reduce duration of COVID-19 symptoms.

**Trial Registration:** ClinicalTrials.gov (NCT04885530).

## INTRODUCTION

Recent clinical trials have studied both novel and repurposed oral therapies for outpatients with mild to moderate coronavirus disease 2019 (COVID-19) without evidence of significant benefit on time to symptom recovery or clinical events.^1–3^ Montelukast, an orally active leukotriene receptor antagonist that inhibits the cysteinyl leukotriene CysLT1 receptor, has anti-inflammatory effects and has been shown to suppress oxidative stress and cytokine production.^4^ While montelukast is currently approved for the treatment of asthma and allergic rhinitis, *in silico* screening based on *in vitro* studies for other RNA viruses supports the plausibility of antiviral activity through inhibition of SARS-CoV-2 protease and polymerase enzymes.^5^ Montelukast may also ameliorate extra-pulmonary manifestations of COVID-19 either directly through blocking of cysteinyl leukotriene receptors, or indirectly through inhibition of the NF-κB signaling pathway.^6^ To date, few studies have evaluated the potential role of montelukast. Three prior clinical studies of hospitalized patients suggest a potential benefit of montelukast on improving symptoms, but these studies were small and had design limitations.^7–9^ There are no known trials assessing the potential role of montelukast in outpatients with mild to moderate COVID-19.

The ongoing Accelerating Coronavirus Disease 2019 Therapeutic Interventions and Vaccines (ACTIV-6) platform randomized clinical trial evaluates repurposed medications in the outpatient setting.^10^ For this study, the ACTIV-6 platform evaluated the effect of montelukast on time to sustained recovery or progression to severe disease in non-hospitalized adults with mild to moderate COVID-19.

## METHODS

### Trial Design and Oversight

The design and rationale for ACTIV-6 has been published.^11^ ACTIV-6 is a double-blind randomized placebo-controlled platform trial evaluating repurposed medications for the treatment of outpatients with mild to moderate COVID-19 in the US.^12^ Using a hybrid decentralized approach allowing virtual enrollment, as well as enrollment through diverse healthcare and community settings, ACTIV-6 has achieved broad reach. The complete protocol and statistical analysis plan are provided in online with this article.

The trial protocol was approved by a central institutional review board with review at each site.

Each study participant provided informed consent using electronic consent. An independent data and safety monitoring committee oversaw participant safety, efficacy, and trial conduct.

### Participants

The montelukast arm was open for enrollment from January 27, 2023 through June 23, 2023, during which 104 sites were open. Participants were identified by enrolling sites or by self-referral via the central study telephone call center.

Study eligibility criteria included: age ≥30 years, SARS-CoV-2 infection confirmed with a positive polymerase chain reaction or antigen test (including home-based testing) within the past 10 days, and actively experiencing ≥2 COVID-19 symptoms for ≤7 days from the time of consent (full eligibility criteria in protocol). Symptoms included fatigue, dyspnea, fever, cough, nausea, vomiting, diarrhea, body aches, chills, headache, sore throat, nasal symptoms, and new loss of sense of taste or smell. Individuals were excluded from participation if they had current or recent hospitalization for COVID-19; ongoing or planned participation in other interventional trials for COVID-19; or current or recent use of, known allergy or sensitivity to, or contraindication to montelukast. Receipt of COVID-19 vaccinations or current use of approved or emergency use authorization therapeutics for outpatient treatment of COVID-19 were allowed.

### Randomization

The adaptive platform design of ACTIV-6 allowed study arms to be added or removed based on evolving data. The period of enrollment for montelukast did not overlap with the enrollment period of other active drugs in the platform. Consequently, the randomization process simplified to a 1:1 matched placebo allocation provided by a random number generator with no pooled placebo contribution.

### Interventions

A 14-day supply of either montelukast or matched placebo was dispensed to the participant via home delivery from a centralized pharmacy. Participants were instructed to self-administer oral montelukast at a dose of one 10 mg tablet or matching placebo daily for 14 days. The initial manufacturer of the active drug, Intas Pharmaceuticals Ltd. (Durham, NC), issued a voluntary recall of the drug in February 2023. A replacement product, which was similar in appearance to the original drug, was sourced through the central pharmacy for the remainder of the study.

### Outcome Measures

The primary outcome was time to sustained recovery within 28 days, defined as the time from receipt of drug to the third of 3 consecutive days without COVID-19 symptoms.^10,12^ Participants who died within the follow-up period were deemed to have not recovered, regardless of whether they were symptom-free for 3 consecutive days. Secondary outcomes included 3 time-to-event endpoints administratively censored at day 28 (number of events permitting): time to death, time to hospitalization or death, or time to first healthcare utilization (a composite of urgent care visits, emergency department visits, hospitalization, or death). Additional secondary outcomes included mean time spent unwell through day 14 and the WHO COVID Clinical Progression Scale on days 7, 14, and 28. Quality of life measures using the PROMIS-29 are being collected through day 180 and are not included in this report.

### Trial Procedures

The ACTIV-6 platform was designed to be conducted remotely, with all screening and eligibility procedures reported by participants and confirmed at the site level. Positive laboratory results for SARS-CoV-2 were verified by study staff prior to randomization. Participants self-reported demographic information, medical history, use of concomitant medications, COVID-19 symptoms, and completed quality of life surveys.

A centralized investigational pharmacy packaged and provided active or placebo study products via courier to the address provided by participants. On February 23, 2023, the ACTIV-6 study team was notified of a voluntary recall of a batch of montelukast by the manufacturer. Although the recall was voluntary, with an abundance of caution, enrollment was paused and distribution of study drug/placebo ceased. By February 27, 2023, a replacement product had been sourced, which matched the original in appearance apart from debossing. Details about drug appearance were removed from the protocol to minimize risk of unblinding, and the study arm was reopened on March 3, 2023. Notification of the recall was sent to the 149 participants who were either currently taking study drug or to whom study drug was in the process of being shipped and who would have been eligible for inclusion in the modified intention-to-treat (mITT) analysis cohort. While these participants were told that their study medication was considered safe, adherence was expected to be influenced by the communication, so the ACTIV-6 investigators decided *a priori* to exclude all notified participants from assessment of the primary and secondary outcomes, but to include these participants in any analyses that adjust for adherence. The target recruitment was increased to achieve a minimum of 1200 participants in the mITT analysis set.

Daily assessments were reported by participants via the study portal during the first 14 days of the study, regardless of symptom status. If participants were not recovered by day 14, the daily assessments continued until sustained recovery or day 28. Planned remote follow-up visits occurred on days 28, 90, and 120. Additional study procedure details are provided in the protocol.

### Statistical Analysis

Proportional hazard regression was used for the time-to-event analysis, and cumulative probability ordinal regression models were utilized for ordinal outcomes. Longitudinal ordinal regressions models were used to estimate the differences in mean time spent unwell.

The planned primary endpoint analysis was a Bayesian proportional hazards model. The primary inferential decision-making quantity was the posterior distribution for the treatment assignment hazard ratio (HR), with a HR >1 indicating a beneficial effect. If the posterior probability of benefit exceeded 0.95 during interim or final analyses, intervention efficacy would be met. To preserve type I error <0.05, the prior for the treatment effect parameter on the log relative hazard scale was a normal distribution centered at 0 and scaled to a standard deviation (SD) of 0.1. All other parameter priors were weakly informative, using the default of 2.5 times the ratio of the SD of the outcome divided by the SD of the predictor variable. The study was designed to have 80% power to detect a HR of 1.2 in the primary endpoint from a total sample size of 1200 participants with planned interim analyses at 300, 600, and 900 participants.

The model for the primary endpoint included the following predictor variables: randomization assignment, age, sex, duration of symptoms prior to study drug receipt, calendar time, vaccination status, geographical location, call center indicator, and baseline symptom severity. The proportional hazard assumption of the primary endpoint was evaluated by generating visual diagnostics such as log-log plots and plots of time-dependent regression coefficients for each model predictor.

Secondary endpoints were analyzed with Bayesian regression models (either proportional hazards or proportional odds). Weakly informative priors were used for all parameters. Secondary endpoints were not used for formal decision-making, and no decision threshold was selected. With the exception of time unwell, the same covariates used in the primary endpoint model were used in the secondary endpoints analyses, provided that the endpoint accrued sufficient events to be analyzed with covariate adjustment.

All available data were utilized to compare each active study drug vs placebo control, regardless of post-randomization adherence. The mITT cohort comprised all participants who were randomized, who did not withdraw before delivery of study drug, and for whom the courier confirmed drug delivery. Study day 1 was defined as the day of study drug delivery. Participants who opted to discontinue data collection were censored at the time of last contact, including those participants who did not complete any surveys or phone calls after receipt of study drug. Missing data among covariates for both primary and secondary analyses were addressed with conditional mean imputations.

A predefined analysis examined potential variations in treatment effects based on participant characteristics. The assessment of treatment effect heterogeneity encompassed age, symptom duration, body mass index (BMI), symptom severity on day 1, calendar time (indicative of circulating SARS-CoV- 2 variants), sex, and vaccination status. Continuous variables were analyzed as such, without stratifying into subgroups. *A priori*, there was concern that the call center could enroll a different and larger population than sites, and an indicator for call center was specified in the model with the intent to assess heterogeneity of treatment effect by site should this occur (see statistical analysis plan). Post-hoc, it was identified that one site had recruited 573 participants. As a sensitivity analysis, the primary endpoint model was expanded to include site-indicator variables from all sites, not just the call center. In an additional post-hoc sensitivity analysis, the baseline hazard of recovery was stratified by site (with sites contributing fewer than 11 participants grouped together). Lastly, the possibility of a differential treatment effect by site was assessed.

Analyses were performed with R^13^ version 4.3 with the following primary packages: rstanarm,^14,15^ rmsb,^16^ and survival.^17^

## RESULTS

### Study Population

Of 1453 participants enrolled in the montelukast study arm, the mITT cohort included 1250 participants who were randomized, received study drug, did not withdraw from the study before receiving study drug, and were not notified of the medication recall. There were 628 participants assigned to receive montelukast and 622 assigned to receive placebo (**Figure 1**).

**Figure 1.**
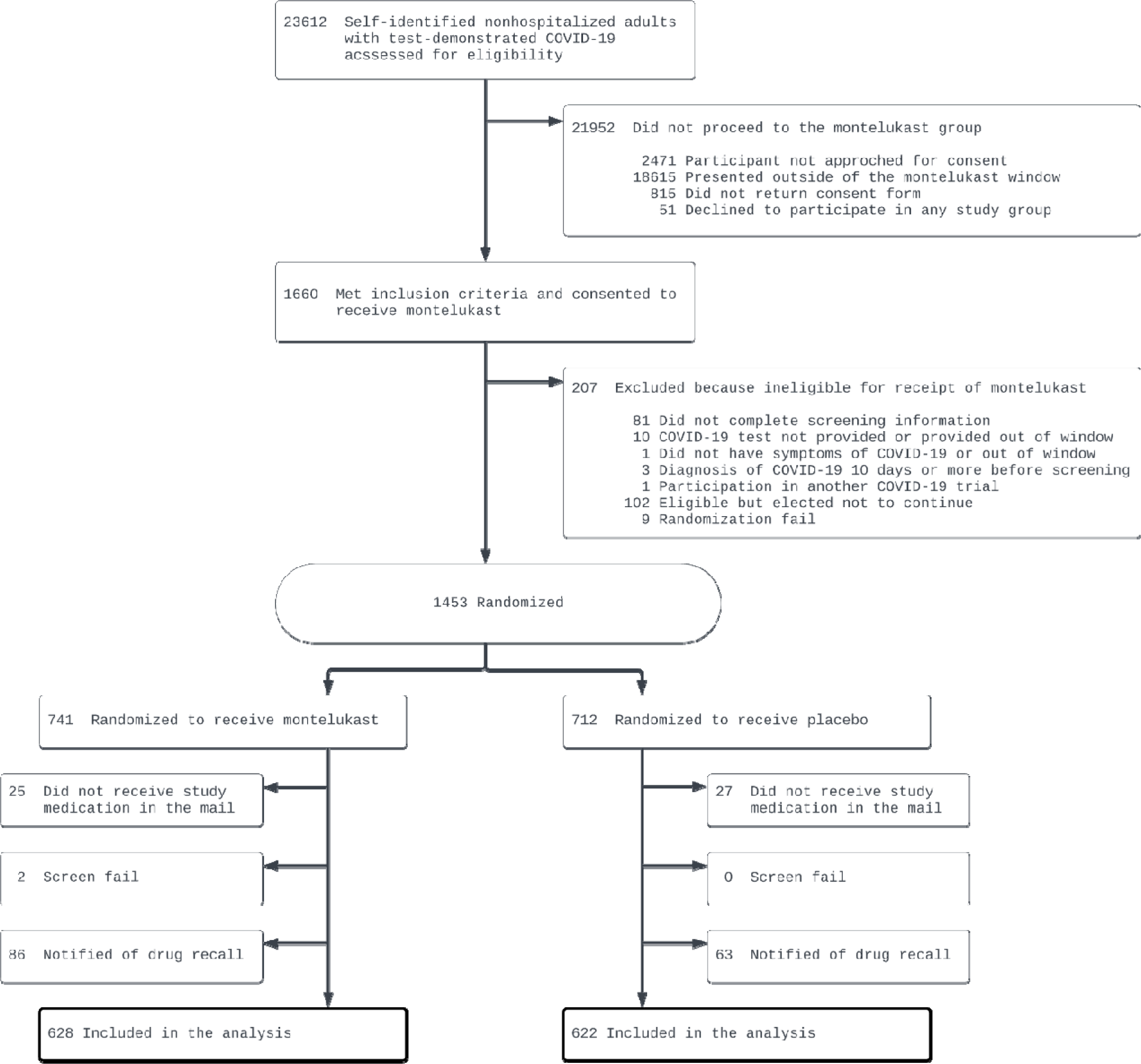
Participant flow in a trial of montelukast for mild to moderate COVID-19

The median age was 53 years (interquartile range [IQR] 42–62); 60.2% were female; most commonly self-reported races were White (78.2%), Black/African American (12.8%), Asian (3.6%), or Middle Eastern (1.5%); and 64.6% of participants identified as Hispanic/Latino. The most common comorbidities were obesity (45.4%) and hypertension (23.1%). Overall, 56.3% of participants reported having received ≥2 SARS-CoV-2 vaccinations and 12.2% of participants reported taking a COVID-19 therapy (**Table 1**).

**Table 1.** Baseline characteristics.

| Variable | Montelukast<br>(n=628) | Placebo<br>(n=622) |
| --- | --- | --- |
| Age, median (IQR), yrs | 52.0 (42.0-61.0) | 54.0 (43.0-62.0) |
| Age < 50 yrs, no. (%) | 278 (44.3) | 241 (38.7) |
| Sex<sup>a</sup>, no. (%) |  |  |
| Female | 388 (61.78) | 365 (58.68) |
| Male | 240 (38.22) | 257 (41.32) |
| Race<sup>b</sup>, not mutually exclusive, no. (%) |  |  |
| American Indian or Alaska Native | 3 (0.48) | 3 (0.48) |
| Asian | 23 (3.66) | 22 (3.54) |
| Black or African American | 85 (13.54) | 75 (12.06) |
| Middle Eastern or North African | 8 (1.27) | 11 (1.77) |
| Native Hawaiian or Other Pacific Islander | 2 (0.32) | 1 (0.16) |
| White | 488 (77.71) | 490 (78.78) |
| None of the above | 16 (2.55) | 19 (3.05) |
| Prefer not to answer | 7 (1.11) | 6 (0.96) |
| Ethnicity, No. (%) |  |  |
| Hispanic/Latino | 415 (66.08) | 392 (63.02) |
| Not Hispanic/Latino | 213 (33.92) | 230 (36.98) |
| Region<sup>c</sup>, No. (%) |  |  |
| Midwest | 83 (13.22) | 98 (15.76) |
| Northeast | 30 (4.78) | 22 (3.54) |
| South | 491 (78.18) | 465 (74.76) |
| West | 24 (3.82) | 37 (5.95) |
| Recruited via call center<sup>d</sup>, no. (%) | 4 (0.64) | 4 (0.64) |
| Body mass index (BMI), median (IQR), kg/m<sup>2</sup> | 29.7 (26.6-33.2) [n=627] | 29.3 (26.4-32.5) |
| BMI > 30, kg/m<sup>2</sup>, no. (%) | 291 (46.3) | 276 (44.4) |
| Weight, median (IQR), kg | 81.6 (74.8-92.1) | 82.3 (72.6-92.0) |
| Medical history<sup>e</sup>, no./total (%) |  |  |
| High blood pressure | 138/603 (22.89) | 139/594 (23.40) |
| Diabetes | 46/603 (7.63) | 68/593 (11.47) |
| Smoker, past year | 51/603 (8.46) | 49/594 (8.25) |
| Asthma | 50/601 (8.32) | 48/593 (8.09) |
| Heart disease | 14/601 (2.33) | 24/593 (4.05) |
| Malignant cancer, No./total (%) | 14/597 (2.35) | 10/588 (1.70) |
| COPD | 10/603 (1.66) | 6/593 (1.01) |
| Chronic kidney disease | 4/602 (0.66) | 6/593 (1.01) |
| SARS-CoV-2 Vaccine status, no. (%) |  |  |
| Not vaccinated | 279 (44.43) | 267 (42.93) |
| Vaccinated, 1 dose | 0 (0.00) | 0 (0.00) |
| Vaccinated, 2+ doses | 349 (55.57) | 355 (57.07) |
| Days between symptom onset and receipt of drug, median (IQR) | 5 (4-6) | 5 (4-6) [n=621] |
| Days between symptom onset and enrollment, median (IQR) | 4 (2-5) [n=619] | 4 (2-5) [n=605] |
| Symptom burden on study day 1<sup>f</sup>, no./No. (%) |  |  |
| None | 21/558 (3.8) | 20/553 (3.6) |
| Mild | 186/558 (33.3) | 183/553 (33.1) |
| Moderate | 341/558 (61.1) | 337/553 (60.9) |
| Severe | 10/558 (1.8) | 13/553 (2.4) |
| COVID-19 medications, no. (%) |  |  |
| Remdesivir | 5 (0.80) | 2 (0.32) |
| Nirmatrelvir+ ritonavir | 64 (10.19) | 64 (10.29) |
| Monoclonal antibodies | 5 (0.80) | 4 (0.64) |
| Molnupiravir | 5 (0.80) | 4 (0.64) |

Abbreviations: BMI, body mass index (calculated as weight in kilograms divided by height in meters squared); COPD, chronic obstructive pulmonary disease; IQR, interquartile range
<sup>a</sup>Participants also had the option to select “unknown,” “undifferentiated,” or “prefer not to answer.” Only “male” and “female” were selected in this cohort.
<sup>b</sup>Participants may have selected any combination of the race descriptors, including “prefer not to answer.” Consequently, the sum of counts over all categories will not match the column total.
<sup>c</sup>The following state groups define each region. Northeast includes Connecticut, Maine, Massachusetts, New Hampshire, Rhode Island, Vermont, New Jersey, New York and Pennsylvania; Midwest includes Indiana, Illinois, Michigan, Ohio, Wisconsin, Iowa, Kansas, Minnesota, Missouri, Nebraska, North Dakota, and South Dakota; South includes Delaware, District of Columbia, Florida, Georgia, Maryland, North Carolina, South Carolina, Virginia, West Virginia, Alabama, Kentucky, Mississippi, Tennessee, Arkansas, Louisiana, Oklahoma, and Texas; West includes Arizona, Colorado, Idaho, New Mexico, Montana, Utah, Nevada, Wyoming, Alaska, California, Hawaii, Oregon, and Washington.
<sup>d</sup>Patients may have alternatively been recruited at local clinical sites.
<sup>e</sup>Medical history was provided by participants, responding to the prompts: “Has a doctor told you that you have any of the following?” and “Have you ever experienced any of the following (select all that apply)” and “Have you ever smoked tobacco products?”
<sup>f</sup>Each day, participants were asked to “Please choose the response that best describes the severity of your COVID-19 symptoms today” with the response options being “no symptoms,” “mild,” “moderate,” and “severe.”

On study day 1, 3.7% of participants reported no symptoms, while the majority reported mild (33.2%) or moderate (61.0%) symptoms. Baseline symptom burden for the 13 COVID-19-related symptoms is reported in **eTable 1**. Participants were enrolled within a median of 4 days of reported symptom onset (IQR 2–5 days) and study drug was delivered within a median of 5 days from symptom onset (IQR 4–6 days) (**eFigure 1)**.

### Primary Outcome

Differences in time to sustained recovery were not observed in either unadjusted Kaplan-Meier curves (**Figure 2**) or covariate-adjusted regression models (**Table 2**). The median time to sustained recovery was 10 days (95% confidence interval [CI] 10–11 days) in both the montelukast and placebo groups. The posterior probability for benefit was 0.63, with an HR of 1.02 (95% credible interval [CrI] 0.92–1.12) (**Figure 3**). Sensitivity analyses yielded similar estimates of the treatment effect (**eFigure 2**).

**Figure 2.**
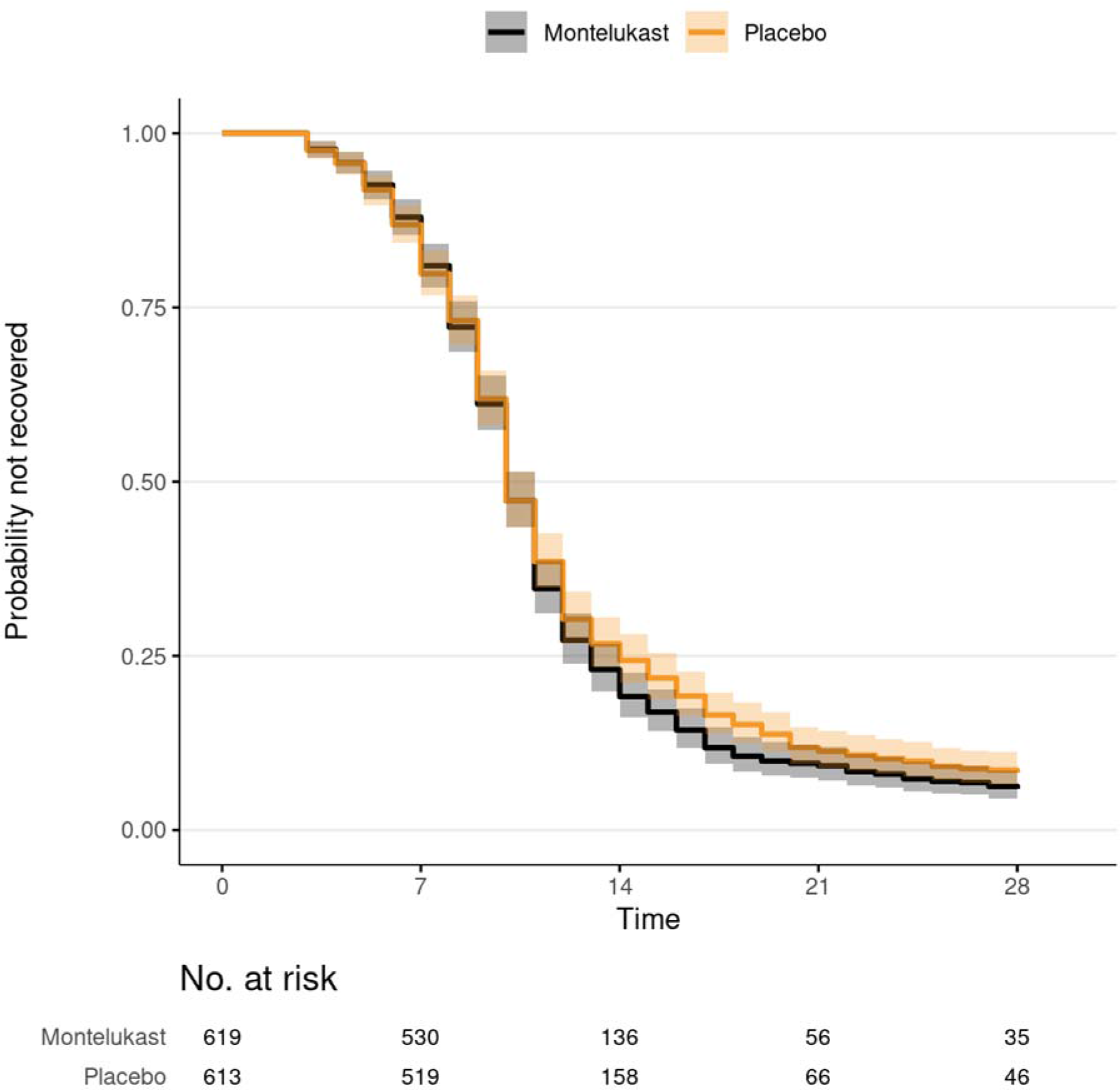
Primary outcome of time to sustained recovery Sustained recovery was defined as the third of 3 consecutive days without symptoms. Eighteen participants were censored for complete nonresponse, 28 were censored after partial response, and all others were followed up until recovery, death, or the end of short-term 28-day follow-up. Median time to sustained recovery was 10 days (95% CI 10-11) in both groups. Shaded regions denote the pointwise 95% CIs.

**Figure 3.**
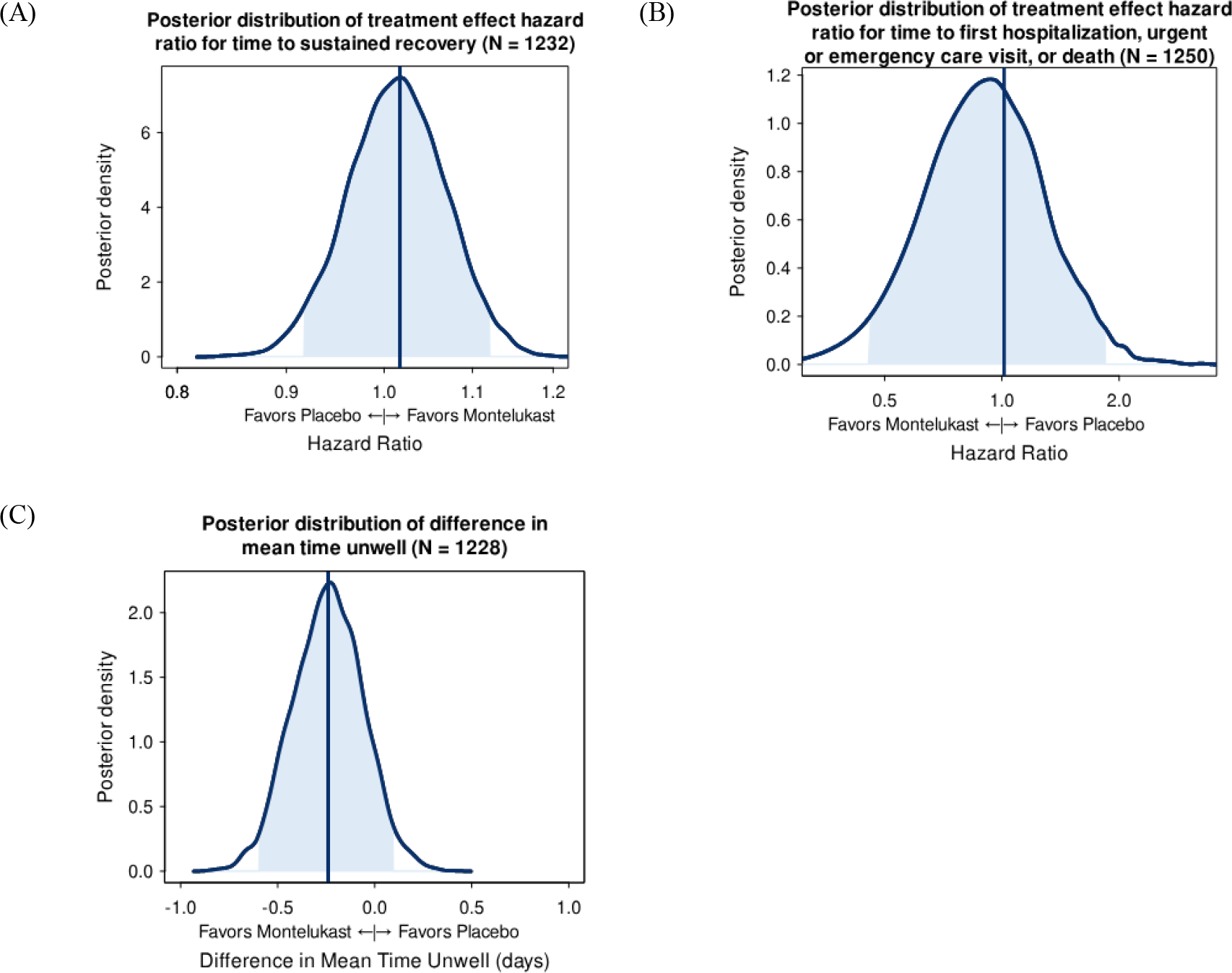
Posterior distributions of treatment effects The vertical lines represent the estimated mean of the posterior distribution. Posterior density is the relative likelihood of posterior probability distribution. Outcomes with higher posterior density are more likely than outcomes with lower posterior density. Blue density lines are kernel density estimates constructed from posterior draws. The posterior density plots of all the covariates in the primary outcome model are shown in the supplement (**e**Figure 10).

**Table 2.**
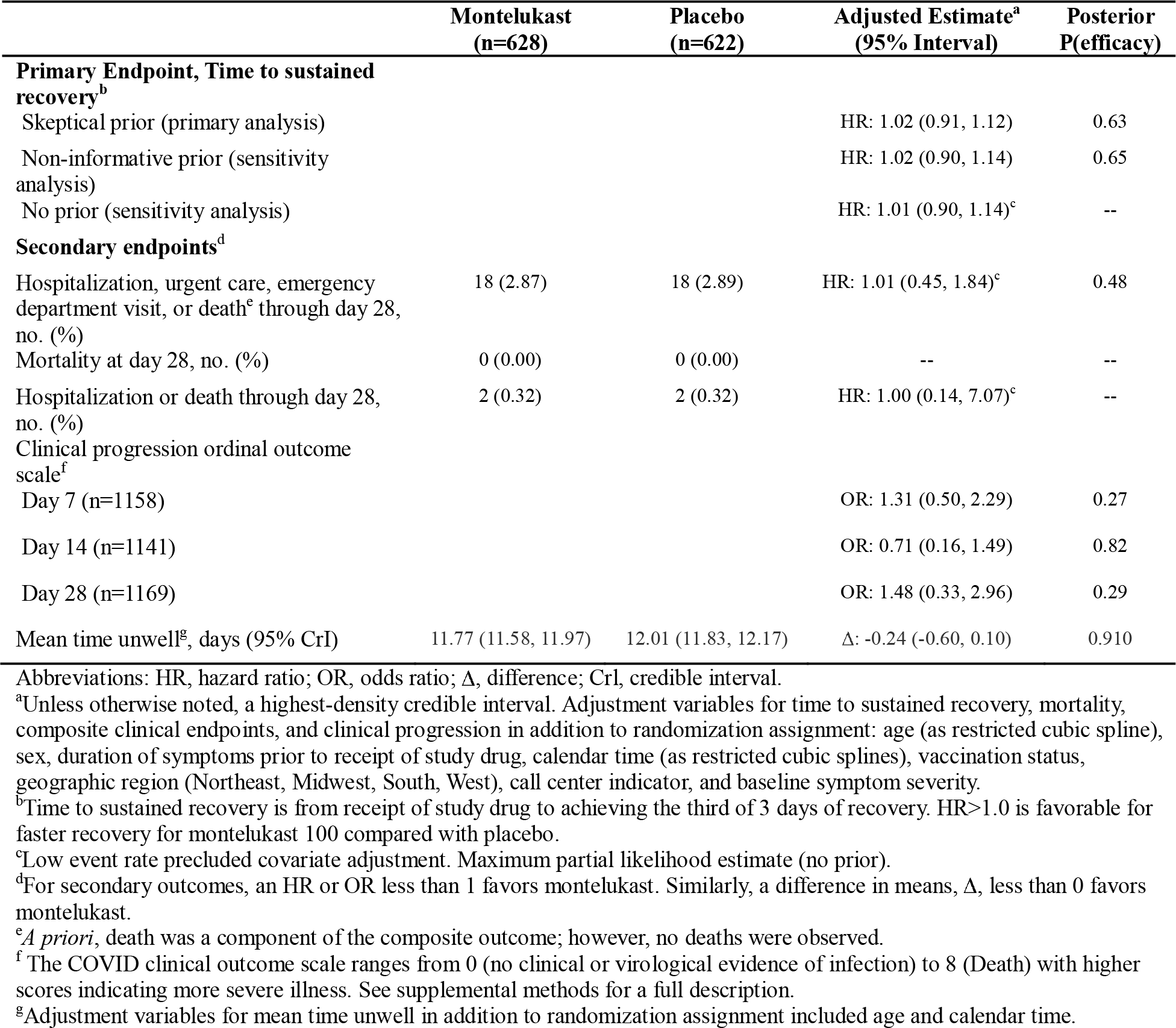
Primary and secondary outcomes.

|  | Montelukast<br>(n=628) | Placebo<br>(n=622) | Adjusted Estimate<sup>a</sup><br>(95% Interval) | Posterior<br>P(efficacy) |
| --- | --- | --- | --- | --- |
| **Primary Endpoint, Time to sustained recovery<sup>b</sup>** |  |  |  |  |
| Skeptical prior (primary analysis) |  |  | HR: 1.02 (0.91, 1.12) | 0.63 |
| Non-informative prior (sensitivity analysis) |  |  | HR: 1.02 (0.90, 1.14) | 0.65 |
| No prior (sensitivity analysis) |  |  | HR: 1.01 (0.90, 1.14)<sup>c</sup> | -- |
| **Secondary endpoints<sup>d</sup>** |  |  |  |  |
| Hospitalization, urgent care, emergency department visit, or death<sup>e</sup> through day 28, no. (%) | 18 (2.87) | 18 (2.89) | HR: 1.01 (0.45, 1.84)<sup>c</sup> | 0.48 |
| Mortality at day 28, no. (%) | 0 (0.00) | 0 (0.00) | -- | -- |
| Hospitalization or death through day 28, no. (%) | 2 (0.32) | 2 (0.32) | HR: 1.00 (0.14, 7.07)<sup>c</sup> | -- |
| **Clinical progression ordinal outcome scale<sup>f</sup>** |  |  |  |  |
| Day 7 (n=1158) |  |  | OR: 1.31 (0.50, 2.29) | 0.27 |
| Day 14 (n=1141) |  |  | OR: 0.71 (0.16, 1.49) | 0.82 |
| Day 28 (n=1169) |  |  | OR: 1.48 (0.33, 2.96) | 0.29 |
| Mean time unwell<sup>g</sup>, days (95% CrI) | 11.77 (11.58, 11.97) | 12.01 (11.83, 12.17) | Δ: -0.24 (-0.60, 0.10) | 0.910 |

Abbreviations: HR, hazard ratio; OR, odds ratio; Δ, difference; CrI, credible interval.
<sup>a</sup>Unless otherwise noted, a highest-density credible interval. Adjustment variables for time to sustained recovery, mortality, composite clinical endpoints, and clinical progression in addition to randomization assignment: age (as restricted cubic spline), sex, duration of symptoms prior to receipt of study drug, calendar time (as restricted cubic splines), vaccination status, geographic region (Northeast, Midwest, South, West), call center indicator, and baseline symptom severity.
<sup>b</sup>Time to sustained recovery is from receipt of study drug to achieving the third of 3 days of recovery. HR>1.0 is favorable for faster recovery for montelukast 100 compared with placebo.
<sup>c</sup>Low event rate precluded covariate adjustment. Maximum partial likelihood estimate (no prior).
<sup>d</sup>For secondary outcomes, an HR or OR less than 1 favors montelukast. Similarly, a difference in means, Δ, less than 0 favors montelukast.
<sup>e</sup>*A priori*, death was a component of the composite outcome; however, no deaths were observed.
<sup>f</sup>The COVID clinical outcome scale ranges from 0 (no clinical or virological evidence of infection) to 8 (Death) with higher scores indicating more severe illness. See supplemental methods for a full description.
<sup>g</sup>Adjustment variables for mean time unwell in addition to randomization assignment included age and calendar time.

### Secondary Outcomes

No deaths occurred; 2 participants in each study group were hospitalized (**Table 2, eFigure 3**). There were 18 (2.9%) participants in the montelukast group and 18 (2.9%) in the placebo group who reported hospital admission or emergency department or urgent care visits (**Table 2**, **eFigure 4**). The HR for the composite healthcare outcome was 1.01 (95% CrI 0.45–1.84) with a posterior probability of efficacy of 0.48 (**Figure 3**).

With clinical events like hospitalization and death being rare among participants, the COVID clinical progression scale (**Supplemental Appendix**) simplifies to a self-reported evaluation of home activity levels (limited vs not) collected on study days 7, 14, and 28 (**eFigure 5**). By day 7, 89.8% of those receiving montelukast and 89.6% of those receiving placebo reported no limitations in activity, not meeting the prespecified thresholds for a beneficial treatment effect (odds ratio [OR] 1.31; 95% CrI 0.50–2.29; P(efficacy)=0.27). Likewise, the difference in mean time unwell was similar between the montelukast and placebo groups (11.8 days [95% CI 11.6–12.0] vs 12.0 days [95% CI 11.8–12.2]; difference -0.24 (95% CrI -0.60 to 0.10; P(days of benefit >0)=0.91; P(days of benefit>1)<0.001) (**Figure 3**).

### Adverse Events and Tolerability

Of the 628 participants assigned to montelukast, 28 did not report taking their medication at least once. Among the 622 assigned to placebo, 32 did not report taking their study medication at least once. Five participants experienced 1 serious adverse event, all among those who reported taking their study drug (**eTable 2**). The 3 events reported in the montelukast group were pneumonia, lower extremity cellulitis, and ovarian torsion. The 2 serious adverse events reported in those randomized to placebo were pneumonia and acute appendicitis. *A priori*, neuropsychiatric events were identified as being of special interest, but no such events were observed.

### Heterogeneity of Treatment Effect Analyses

Analyses of *a priori* defined characteristics found that as time from symptom onset to receipt of study drug increased beyond 9 days, the treatment effect favored placebo (**eFigure 6**), but this represented less than 2% of study participants. Similarly, the treatment effect in participants no longer reporting symptoms on study day 1 favored placebo, but this represented just 41 participants (**eFigure 7**). **eFigures 8 and 9** show that the main results and treatment effect were not influenced by site. No other factors were associated with the treatment effect.

## DISCUSSION

In this randomized trial of 1250 adults with mild to moderate COVID-19, montelukast 10 mg daily for 14 days did not improve time to sustained recovery compared with placebo. Several recent studies have suggested a possible benefit from montelukast for inpatients with COVID-19, but these studies all had design limitations. In one open-label clinical trial, 180 hospitalized patients with moderate to severe COVID-19 were randomized to 1 of 3 arms: gabapentin, gabapentin plus montelukast (10 mg daily), or dextromethorphan (control)^9^ for 5 days. The authors found that gabapentin plus montelukast reduced the frequency and severity of cough to a greater extent than gabapentin alone; however, the dextromethorphan group performed better than either of these 2 experimental groups. Another clinical trial randomized 180 hospitalized patients with COVID-19 to receive standard of care alone or into 1 of 2 experimental groups: montelukast 10 mg daily or 20 mg daily for 5 days.^8^ The study found that inflammatory markers were significantly lower at day 5 in the montelukast groups compared with standard of care alone; however, only the higher-dose montelukast group had improved pulmonary function testing. Too few clinical events of interest occurred to adequately assess differences between the groups. Finally, a retrospective study of 92 hospitalized patients compared the COVID-19 ordinal scale of 30 patients receiving montelukast to 62 patients not receiving monteleukast.^7^ The authors reported significantly fewer clinical deterioration events (increase in COVID-19 ordinal scale) at day 3 of hospitalization in those receiving montelukast (10% vs 32%; P=0.022). It is possible that these 3 studies found a potential benefit from montelukast for more severe COVID-19, while ACTIV-6 did not find a benefit in patients with less severe disease. However, the design limitations of the prior studies raise concerns that montelukast may not be beneficial for acute COVID-19 in any setting.

## Limitations

Due to the dynamic nature of the pandemic and the characteristics of the enrolled population, there were few clinical events observed in this trial. This limited our ability to adequately evaluate the treatment effect on clinical outcomes, both for the primary and secondary endpoints. A notable constraint of the decentralized trial approach is the necessity to send the study drug via courier, leading to a median delay of 1 additional day between symptom onset and drug receipt. This delay might be significant for a proposed antiviral mechanism of action. We achieved a median time from symptom onset to receiving study drug of 5 days, with 90% of patients obtaining their study drug within a 7-day timeframe, which is a relevant timeframe for people in the outpatient setting who might be seeking treatment. Also, with a distribution of time from symptom onset to drug delivery we were able to include this in the analysis of heterogeneity of treatment effect to inform timing of administration. We excluded persons notified of the medication recall although we do not expect that to have contributed to any imbalance between study groups. Finally, 46% of enrollment occurred at 1 site. There was no evidence of a difference in treatment effect for this site when compared with other sites, and we expect the results generalize broadly to the United States population.

## Conclusions

Among outpatient adults with mild to moderate COVID-19, treatment with montelukast 10 mg daily, compared with placebo, did not shorten time to sustained recovery.

## Data Availability

ACTIV-6 is a platform trial using shared placebos. On completion of the platform trial, when there is no risk of unblinding across study arms, the data will be made publicly available by depositing it in an approved data repository such as NHLBI BioData Catalyst.

## Acknowledgments

We thank Samuel Bozzette, MD, PhD, of the National Center for Advancing Translational Sciences (NCATS), for his role in the trial design and protocol development. We also thank the ACTIV-6 Data Monitoring Committee and Clinical Events Committee Members (listed below) for their contributions. Data Monitoring Committee: Clyde Yancy, MD, MSc, Northwestern University Feinberg School of Medicine; Adaora Adimora, MD, University of North Carolina, Chapel Hill; Susan Ellenberg, PhD, University of Pennsylvania; Kaleab Abebe, PhD, University of Pittsburgh; Arthur Kim, MD, Massachusetts General Hospital; John D. Lantos, MD, Children’s Mercy Hospital; Jennifer Silvey-Cason, Participant representative; Frank Rockhold, PhD, Duke Clinical Research Institute; Sean O’Brien, PhD, Duke Clinical Research Institute; Frank Harrell, PhD, Vanderbilt University Medical Center; Zhen Huang, MS, Duke Clinical Research Institute. Clinical Events Committee: Renato Lopes, MD, PhD, MHS, W. Schuyler Jones, MD, Antonio Gutierrez, MD, Robert Harrison, MD, David Kong, MD, Robert McGarrah, MD, Michelle Kelsey, MD, Konstantin Krychtiuk, MD, Vishal Rao, MD all of the Duke Clinical Research Institute, Duke University School of Medicine. Elizabeth E.S. Cook of the Duke Clinical Research Institute provided editorial support.

## Author Contributions

Drs Rothman, Naggie, Hernandez, and Lindsell had full access to all the blinded data in the study. Dr Stewart was provided curated study data and takes responsibility for the integrity of the data analysis. All authors contributed to the drafting and review of the manuscript and agreed to submit for publication.

